# The epidemiological scenario of leptospirosis in Brazil from 2015 to 2024: An ecological study of socio-environmental and climatic determinants

**DOI:** 10.64898/2026.04.15.26350927

**Authors:** George S. C. Fernandes, Bruna O. P. Azevedo, Deborah K. Damiano, Mikael V. R. Lima, Pablo P. Macena, Aline F. Teixeira, Giovana C. Barazzone, Ana L. T. O. Nascimento, Alexandre P.Y. Lopes

**Affiliations:** Laboratório de Desenvolvimento de Vacinas, Instituto Butantan, Avenida Vital Brasil, São Paulo, SP, Brazil; Programa de Pós-Graduação Interunidades em Biotecnologia, Instituto de Ciências Biomédicas, Universidade de São Paulo, SP, Brazil

## Abstract

**Background:** Leptospirosis is a neglected tropical disease with substantial public health impact in Brazil, closely associated with socio-environmental vulnerabilities and climatic extremes. This study analyzed the epidemiological profile, spatiotemporal distribution, and climatic influences on leptospirosis incidence and lethality in Brazil from 2015 to 2024.

**Methods:** An ecological time-series study was conducted using secondary data from the Notifiable Diseases Information System (SINAN). Variables included geographic region, probable infection environment, occupational, and educational level (ISCED-2011). The spatiotemporal correlation between disease incidence and El Niño-Southern Oscillation (ENSO) anomalies was assessed using the Oceanic Niño Index (ONI) and Spearman’s rank correlation coefficient.

**Results:** A total of 31,397 cases were notified, with an annual average of 3,140 cases. The South and North regions exhibited the highest incidence rates, while the Northeast and Southeast presented lethality rates above the national average (9.20%). A marked reduction in notifications occurred during the COVID-19 pandemic. Contaminations occurred predominantly in the domiciliary environment (64%). Rural workers (27.45%) and civil construction workers (18.63%) were the most affected occupational groups, with a higher incidence among illiterate and low-education populations. Climatic analysis revealed a positive spatial correlation between El Niño intensification and leptospirosis incidence in the South and Southeast, and a negative correlation in specific Northeastern states.

**Conclusion:** The dynamics of leptospirosis in Brazil are complex and multifactorial, strongly influenced by macroclimatic variations and driven by deficits in basic sanitation and urbanization. Mitigating the disease burden requires sustained, region-specific public health strategies, targeted infrastructure improvements, and enhanced epidemiological surveillance to address underreporting.

**Author Summary:** Leptospirosis is a life-threatening, neglected tropical disease that disproportionately affects vulnerable populations in Brazil. Despite its significant impact on public health, the way climatic events and socioeconomic factors interact to drive the disease remains complex and regionally distinct. In this comprehensive 10-year study (2015–2024), we analyzed over 31,000 cases across all Brazilian states. We discovered that the home environment is the primary site of infection, largely due to inadequate basic sanitation. The disease takes a severe toll on individuals with lower educational levels and those engaged in rural or civil construction work. Furthermore, we demonstrated that the El Niño climate phenomenon acts as a major spatiotemporal modulator for leptospirosis, triggering infection spikes in the flood-prone South and Southeast regions, while reducing cases during severe droughts in the Northeast. Our findings underscore that leptospirosis is not merely an infectious disease, but a direct symptom of socio-environmental vulnerability. These insights are crucial for public health policymakers to design localized, climate-informed early warning systems and to prioritize basic sanitation infrastructure where it is most urgently needed.

## INTRODUCTION

Leptospirosis is a zoonotic infection caused by pathogenic species of the genus *Leptospira*, with worldwide distribution and endemic occurrence in both urban and rural areas of developing countries, including Brazil (1). It is considered a neglected infectious disease of significant human and veterinary concern. The prevalence is substantially higher in tropical and subtropical regions than in temperate areas, largely due to the prolonged survival of leptospires in warm, humid environments. Disease outbreaks are commonly associated with the rainy season, particularly during periods of heavy rainfall and flooding, a situation further exacerbated by climate change (2).

The pathogenic group comprises the etiological agents of leptospirosis, whereas the saprophytic group consists of free-living, non-pathogenic organisms. Genetically, leptospires are classified into four groups: P1 (pathogenic), P2 (intermediate), and S1 and S2 (saprophytic) (3). In addition, they are serologically categorized into serogroups and serovars based on the antigenic heterogeneity of surface-exposed lipopolysaccharides (LPS) (4). To date, more than 300 pathogenic serovars have been identified (3, 5). Morphologically, leptospires are characterized by distinctive hooked ends and possess two periplasmic flagella inserted at opposite poles within the periplasmic space, which confer motility (6). Structurally, they exhibit a typical double-membrane architecture in which the cytoplasmic membrane and peptidoglycan layer are closely associated and covered by an outer membrane. The outer membrane contains LPS, which represents the major antigenic component of these bacteria (5, 7).

In urban settings, rodents serve as the primary reservoir of *Leptospira*, as they typically remain asymptomatic while maintaining long-term renal colonization. Infected rodents continuously shed viable leptospires in their urine, contaminating the environment and facilitating transmission through direct contact or indirectly via contaminated soil and water (4, 8, 9). Following exposure through abraded or water-sodden skin or mucous membranes, pathogenic leptospires rapidly penetrate host tissues, overcoming biological barriers and resisting complement-mediated killing. They can disseminate hematogenously and reach target organs, including the kidneys (via the proximal tubules), liver, and lungs, within one hour of infection (4), highlighting their marked invasive capacity (9).

Clinically, leptospirosis typically presents a biphasic course, consisting of an initial septicemic phase followed by an immune phase characterized by antibody production. The disease manifests with a broad spectrum of nonspecific symptoms, often resembling other acute febrile illnesses. Common manifestations include fever, headache, anorexia, myalgia, and gastrointestinal disturbances (10). In severe cases, leptospirosis may progress to Weil’s disease or severe pulmonary hemorrhagic syndrome, with reported mortality rates ranging from 5% to 40% (11).

In addition to its public health relevance, leptospirosis imposes a significant economic burden on the agricultural sector, as infection in livestock can result in abortions, stillbirths, infertility, reduced milk production, and death (4).

Prophylactic measures remain the most effective strategy for controlling leptospirosis, particularly because improvements in sanitation and sustained rodent control are often difficult to implement. Despite extensive research efforts worldwide, a universal vaccine has not yet been achieved. Currently, bacterins, formulated from inactivated whole-cell preparations, are the only commercially available vaccines. These are primarily used in veterinary medicine and, in some countries, are administered to high-risk occupational groups in humans (12-14). However, bacterin-based vaccines have important limitations, including serovar-specific protection and short duration of immunity, necessitating annual booster doses.

Considering the clinical significance of leptospirosis, in the last years several studies have been conducted to understand the epidemiology of this disease in Brazil. However, due to the complexity of the disease and lack of information, many gaps remain. Thus, this study aims to update the epidemiological profile of leptospirosis in Brazil by analyzing spatiotemporal distribution, incidence, and lethality rates from 2015 to 2024. Furthermore, it evaluates the impact of socio-environmental vulnerabilities—such as the probable environment of infection, occupational sector, and educational level—alongside the macroclimatic influence of the El Niño-Southern Oscillation (ENSO) on the disease dynamics. The findings are expected to provide relevant insights into the epidemiology of the infection and its associated risk factors, which could support the implementation of targeted public health interventions and climate-informed early warning systems.

## METHODS

### Study area

An ecological time-series study was conducted using an exploratory approach, encompassing the Brazilian national territory. The investigation period comprised the years from 2015 to 2024. The geographic analysis unit consisted of the 27 Federative Units (FUs), evaluated both individually and grouped into the five major regions (North, Northeast, Central-West, Southeast, and South) (Fig. 1).

**Fig 1.**
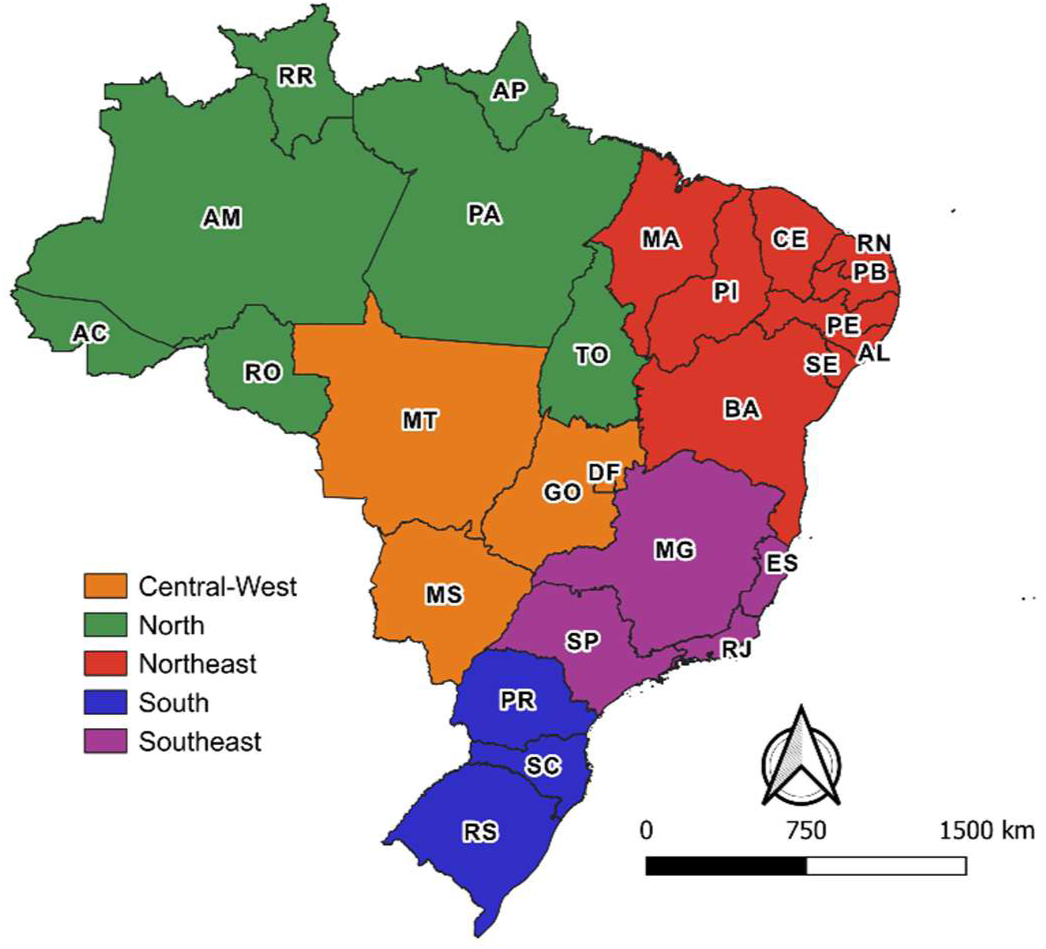
Political map of Brazil stratified by regions. The map illustrates the study area and highlights the grouping of the 27 Federative Units (FUs). The colors indicate the five official regions (North, Northeast, Central-West, Southeast, and South), which served as a spatial scale for aggregating epidemiological data. AC: Acre, AL: Alagoas, AM: Amazonas, AP: Amapá, BA: Bahia, CE: Ceará, DF: Distrito Federal, ES: Espírito Santo, GO: Goiás, MA: Maranhão, MG: Minas Gerais, MS: Mato Grosso do Sul, MT: Mato Grosso, PA: Pará, PB: Paraíba, PE: Pernambuco, PI: Piauí, PR: Paraná, RJ: Rio de Janeiro, RN: Rio Grande do Norte, RO: Rondônia, RR: Roraima, RS: Rio Grande do Sul, SC: Santa Catarina, SE: Sergipe, SP: São Paulo, TO: Tocantins. AC: Acre, AL: Alagoas, AM: Amazonas, AP: Amapá, BA: Bahia, CE: Ceará, DF: Distrito Federal, ES: Espírito Santo, GO: Goiás, MA: Maranhão, MG: Minas Gerais, MS: Mato Grosso do Sul, MT: Mato Grosso, PA: Pará, PB: Paraíba, PE: Pernambuco, PI: Piauí, PR: Paraná, RJ: Rio de Janeiro, RN: Rio Grande do Norte, RO: Rondônia, RR: Roraima, RS: Rio Grande do Sul, SC: Santa Catarina, SE: Sergipe, SP: São Paulo, TO: Tocantins.

### Participants (study population)

The study population comprised all confirmed leptospirosis cases reported in Brazil from 2015 to 2024. Secondary data were extracted from the Brazilian Notifiable Diseases Information System (SINAN), publicly available through the Informatics Department of the Unified Health System (DATASUS) (accessed on September 22, 2025). Data for 2024, updated by the Ministry of Health on July 9, 2025, are preliminary and subject to revision. To calculate incidence rates, annual resident population estimates provided by the Brazilian Institute of Geography and Statistics (IBGE) were used as denominators.

No sample size calculation was performed, as the study considered the census universe of the notifications registered in the official system (intentional non-probabilistic sampling).

### Data collection (quantitative/categorical variables)

The variables of interest were extracted from the SINAN Notification/Investigation Form. Explanatory (independent) variables included the year of notification, Federative Unit of notification, geographic region, area of residence (urban or rural), exposure setting (leisure, domiciliary, or occupational), and educational level. Based on the extracted data, the outcomes of interest (dependent variables) analyzed were the incidence and lethality rates of leptospirosis. Because it is a retrospective study based on secondary data in the public domain, strictly anonymous and without the possibility of individual identification, the present work is exempt from review by a Research Ethics Committee, in accordance with the guidelines of the National Health Council (CNS) and the Brazilian General Data Protection Law (LGPD – Law No. 13,709/2018).

To characterize the occupational profile of the notified cases, the top 100 occupations with the highest frequency reported in the Epidemiological Surveillance Panel of the Brazilian Ministry of Health were collected. This set accounted for 83.18% of the records with the "occupation" variable filled in. The other occupations, due to high dispersion and low individual frequencies, were not detailed in this step, accounting for the remaining 16.82% of the records.

In order to systematize the analysis and facilitate pattern identification, the 100 selected occupations were regrouped into eight categories defined by the nature of the economic activity and the similarity of the work processes.

It should be noted that the relative frequencies presented refer exclusively to cases in which the occupation was filled out in the system of origin. Thus, the percentages presented reflect the distribution among the cases with known occupation, and the possibility of information bias resulting from the non-completion of this variable in the notification forms (ignored/blank) must be considered.

The variable regarding education, originally registered in SINAN according to the Brazilian educational system, was recoded. To ensure international analytical comparability, methodological equivalence was established in accordance with the Operational Manual of the International Standard Classification of Education (ISCED) (15). Consequently, the original strata were aggregated as follows: ISCED 0 (illiteracy or no formal education), ISCED 1-2 (primary education, corresponding to the 1st to 9th grade and incomplete high school), ISCED 3-4 (complete high school, referring to the 1st to 3rd year and incomplete higher education), and ISCED 5-6 (complete technical level or higher education).

The climatic variations associated with the El Niño-Southern Oscillation (ENSO) phenomenon were parameterized using the Relative Oceanic Niño Index (RONI). The monthly historical series, which categorizes the occurrence and magnitude of El Niño, La Niña, and neutral phases, was obtained from the Golden Gate Weather Services repository (Jan Null, CCM) and covers the entire time frame of the study.

In the analytical modeling, the RONI index was operationalized as a continuous variable, restricting the analysis to periods characterized by positive oceanic thermal anomalies. To investigate the potential latency effect of climatic changes on leptospirosis incidence, the data were structured to incorporate and test time lags of one and two months in the analytical models.

The underlying data for the figures and tables are provided in the Supplementary Material.

### Statistical analyses and map preparation

The distribution of the data was assessed for normality. Because the normality criteria were not achieved, nonparametric statistical methods were employed. To quantify the magnitude and direction of the spatial and temporal association between the intensity of positive oceanic anomalies (RONI index) and the leptospirosis incidence rates, Spearman’s rank correlation coefficient (ρ) was employed. All inferential analyses were conducted in a two-tailed manner, with a significance level of 5% (p < 0.05) and 95% confidence intervals. Data processing and statistical analyses were performed in the GraphPad Prism software.

The analysis of the spatial distribution of leptospirosis in the Brazilian territory was represented by choropleth maps. For the definition of the class intervals, a partitioning approach using medians applied to the global dataset (n=270 observations) was adopted, corresponding to the annual incidence rates of all 27 Federative Units over the analyzed decade. The values were arranged in ascending order: the first median divided the total set, isolating the upper half of the data; then, the median of the remaining lower subset was successively calculated, repeating the process to define the limits of the lower-incidence classes. This non-parametric stratification is justified because it allows progressive isolation of extreme values (outliers) in the upper classes, preventing atypical epidemic peaks from distorting the visual representation of the historical series and ensuring a reliable visualization of the country’s basal endemicity scenario. The territorial meshes used in the cartographic preparation were obtained from public-domain geospatial databases of the Brazilian Institute of Geography and Statistics (IBGE). Spatial data processing, attribute table pairing, and final map preparation were conducted using the QGIS software.

## RESULTS

### Overview of leptospirosis cases in Brazil over the 10-year period

Human leptospirosis is a disease that affects all Brazilian states. According to the data collected from 2015 to 2024, about 31,397 cases were notified. The number of annual cases in the country averaged 3140 (Tab 1). Within this period, 2021 had the lowest number of registered cases (1744), while 2015 had the highest, with 4337 notifications (Tab 2). Over these 10 years, the Central-West region was the least affected by the disease, with an average of 64 annual cases and a lower variation of cases among the years.

**Tab 1.** Average of the epidemiological indicators of leptospirosis in Brazil (2015–2024).

| Study Areas | Abbr. | Demographic<br>Density<br>(Inh./Km<sup>2</sup>) | Annual<br>Mean Cases | Incidence<br>(cases/100.000<br>Inh.) | Lethality (%) |
| --- | --- | --- | --- | --- | --- |
| Brazil | BR | 24.77 | 3140 | 1.49 | 9.20 |
| Central-West |  | 10.20 | 64 | 0.39 | 7.89 |
| Distrito Federal | DF | 526.28 | 22 | 0.72 | 11.39 |
| Goiás | GO | 20.75 | 20 | 0.28 | 6.73 |
| Mato Grosso | MG | 3.88 | 15 | 0.41 | 10.84 |
| Mato Grosso do Sul | MS | 7.82 | 9 | 0.31 | 3.16 |
| North |  | 4.81 | 494 | 2.71 | 6.04 |
| Acre | AC | 5.41 | 242 | 28.03 | 0.67 |
| Amapá | AP | 6.00 | 52 | 6.20 | 5.20 |
| Amazonas | AM | 2.68 | 49 | 1.18 | 11.29 |
| Pará | PA | 6.94 | 111 | 1.28 | 14.43 |
| Rondônia | RO | 7.51 | 30 | 1.71 | 6.36 |
| Roraima | RR | 2.73 | 3 | 0.39 | 1.85 |
| Tocantins | TO | 5.70 | 8 | 0.49 | 0.71 |
| Northeast |  | 36.85 | 522 | 0.91 | 14.12 |
| Alagoas | AL | 120.13 | 47 | 1.41 | 13.20 |
| Bahia | BA | 26.37 | 96 | 0.65 | 14.60 |
| Ceará | CE | 61.48 | 65 | 0.70 | 12.18 |
| Maranhão | MA | 21.52 | 26 | 0.37 | 16.38 |
| Paraíba | PB | 71.32 | 18 | 0.43 | 12.50 |
| Pernambuco | PE | 97.72 | 222 | 2.30 | 14.56 |
| Piauí | PI | 13.01 | 8 | 0.23 | 0.00 |
| Rio Grande do Norte | RN | 66.62 | 13 | 0.36 | 14.50 |
| Sergipe | SE | 105.17 | 29 | 1.24 | 21.34 |
| South |  | 52.12 | 1141 | 3.80 | 5.66 |
| Paraná | PR | 57.54 | 349 | 3.05 | 8.09 |
| Rio Grande do Sul | RS | 40.44 | 508 | 4.45 | 4.92 |
| Santa Catarina | SC | 75.24 | 284 | 3.98 | 4.24 |
| Southeast |  | 95.85 | 919 | 1.04 | 12.79 |
| Espírito Santo | ES | 87.65 | 63 | 0.95 | 3.15 |
| Minas Gerais | MG | 36.17 | 167 | 0.79 | 7.81 |
| Rio de Janeiro | RJ | 395.48 | 214 | 1.23 | 15.46 |
| São Paulo | SP | 185.58 | 500 | 1.09 | 14.49 |

It presents the annual average of cases, incidence rate (per 100,000 inh), and lethality (%) by region, Federative Unit, and country, along with the respective population density. Abbr.: State abbreviation; Inh.: Inhabitants.

**Tab 2.** Absolute number of confirmed leptospirosis cases in Brazil and regions.

| Leptospirosis Cases in Brazilian Regions per Year |  |  |  |  |  |  |
| --- | --- | --- | --- | --- | --- | --- |
| Year | Brazil | Central-West | North | Northeast | South | Southeast |
| 2015 | 4337 | 80 | 1312 | 424 | 1573 | 948 |
| 2016 | 3065 | 74 | 480 | 324 | 1214 | 973 |
| 2017 | 3014 | 54 | 516 | 463 | 1066 | 915 |
| 2018 | 3079 | 70 | 495 | 470 | 1023 | 1021 |
| 2019 | 3698 | 78 | 491 | 637 | 1405 | 1087 |
| 2020 | 1833 | 57 | 307 | 273 | 531 | 665 |
| 2021 | 1744 | 36 | 288 | 324 | 543 | 553 |
| 2022 | 3194 | 52 | 319 | 1000 | 797 | 1026 |
| 2023 | 3343 | 61 | 364 | 650 | 1229 | 1039 |
| 2024 | 4090 | 81 | 367 | 654 | 2025 | 963 |

The data detail the annual notifications graphically represented in Fig 2.

The North region presented an average of 494 annual cases, while the Northeast had 522 registered cases. These numbers, although close, diverge strongly due to the regions’ demographic density: the North region has 4.81 inhabitants per km^2^, while the Northeast has 36.85 inhabitants per km^2^ (Tab 1). The most populous regions of the country, the Southeast (95.85 inh/km^2^) and the South (52.12 inh/km^2^), harbor most of the cases, with 919 and 1141, respectively.

When evaluating incidence, which translates into epidemiological risk, the north region follows the south region, which has the second-highest population density in the country. The highest incidence rate of the disease was in the state of Acre, with about 28.03 cases/100,000 Inh, a number significantly higher than in any other Brazilian state. Rio Grande do Sul ranked second, with an incidence rate of 4.45 cases/100,000 Inh, followed by Santa Catarina and Paraná states, with 3.98 and 3.05 cases/100,000 Inh, respectively. The lowest incidence rate was observed in the state of Piauí in the Northeast, with about 0.23 cases/100,000 Inh. In contrast, the state of Pernambuco presented a high index (2.30 cases/100,000 Inh), compared to the other states in the region. The Southeast presents an overall incidence of 1.04 cases/100,000 inh, with higher prevalence in São Paulo and Rio de Janeiro (Tab 1).

The lethality rate, a direct indicator of the clinical severity and disease outcome, was 9.20% in Brazil during the study period. However, a regional variation was observed, with the northeast and southeast regions standing out for presenting lethality rates above the national average. In the overall average, the Northeast registered a lethality of 14.12%, with Sergipe (21.34%), Maranhão (16.38%), Bahia (14.60%), Pernambuco (14.56%), and Rio Grande do Norte (14.50%) standing out for having lethality above the average. It is interesting to note that the lethality rate in Sergipe is notably high, particularly when compared with data from the Pernambuco region, which has a lower incidence of cases. (Tab 1). The Southeast region had an average lethality of 12.79%, with São Paulo and Rio de Janeiro presenting the highest lethality rates. The Central-West (7.89%), North (6.04%), and South (5.66%) regions registered the lowest indices. Although the Central-West region has a lower lethality than other regions, the state of Mato Grosso and the Federal District stand out for their low incidence and a lethality rate above the overall average. In contrast, in the north region, the state of Acre stands out for its very high incidence and a lethality rate of 0.67% (Tab 1).

### Distribution of leptospirosis cases by region of Brazil

According to the distribution of cases over the 10 years comprised in the study, the South region began the last decade with 1573 cases in the year 2015 and followed a downward trend until 2018, when it reached 1023 cases, accompanying the national average, 4337 in 2015 and 3079 in 2018 (Fig 2 and Tab 2). From 2022 onward, a sharp increase in cases was observed, peaking in 2024 with 2,025 cases, the highest number on record. This surge was exacerbated by severe flooding in the state of Rio Grande do Sul. In the Southeast region, 948 cases were reported in 2015, followed by a gradual upward trend that reached a peak of 1,087 cases in 2019, representing a 15% increase compared to the region’s annual average of 919 cases.

**Fig 2.**
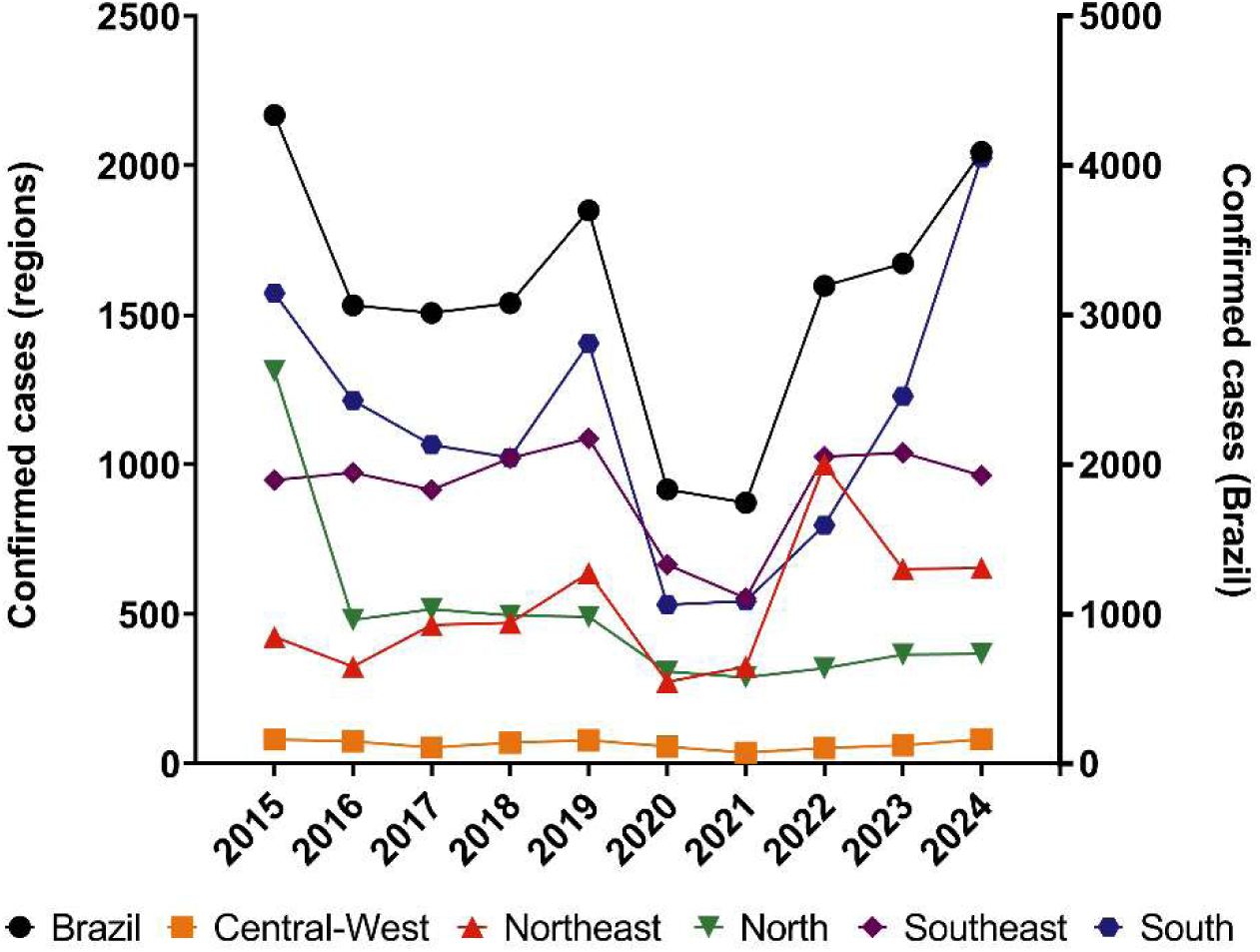
Historical series of confirmed leptospirosis cases in Brazil and regions. The left vertical axis shows the annual number of cases for each region of the country, while the right vertical axis shows the consolidated total of cases across the national territory (black line).

In the year 2015, the North region presented 1312 cases, far above its annual average of 494 cases, due to the numbers of the state of Acre, with an exceptional 601 cases, when the state faced an environmental disaster, with a flood considered the largest in history (the Acre river registered a record on March 4, affecting more than 100,000 people) (Fig 2 and Tab 2). From 2016 onwards, the number of cases remained close to the annual average (480 cases), with minimal variation among the other periods. In 2020 and 2021, a reduction in notifications was observed, but not as pronounced as in the south and southeast regions. Interestingly, in 2024, the year of the second-largest flood in the state of Acre, no significant increase in notifications was observed, ending the analyzed series with 367 notified cases. The Central-West region presented the lowest notification index during the entire analyzed period, with a reduction in the number of cases being observed only in 2021 (Tab 2).

The Northeast region began the period with 424 cases in 2015. A reduction in the number of cases was observed in 2016, and from the following year onwards, reaching 637 cases, above the region’s annual average of 522 cases. Notably, an isolated peak was observed in 2022 due to catastrophic rains in several states of the region. In this period, 1,000 cases were identified, momentarily dissociating from the stability trend observed in previous and subsequent years. A reduction was observed, with 650 cases in 2023 and 654 cases in 2024 (Tab 2).

In a general manner in the studied regions, in the 2020-2021 biennium, a "U"-shaped pattern of descent and ascent. The generalized decline coincides with the critical period of the COVID-19 pandemic, suggesting a possible influence of underreporting or social isolation measures on the transmission dynamics. From 2022 onwards, the number of cases returned to the indices observed in the previous period.

### Annual incidence of leptospirosis cases in Brazil

The pattern of the annual distribution of incidence follows the annual distribution of the number of cases (Fig 3). It is observed that the North region leads the incidence number in 2015 (7.51 cases/100,000 Inh), the highest recorded during the 10 years analyzed, followed by a sharp drop in subsequent years (Tab 3). The South of the country, in turn, stood out for presenting the highest incidence rates throughout the evaluated decade, with expressive growth, especially between the years 2021 (1.79 cases/100,000 Inh) and 2024 (6.53 cases/100,000 Inh). The Southeast and Northeast Regions maintained incidence rates below the annual national average (1.49 cases/100,000 Inh) during most of the analyzed period. It is highlighted, however, that the Northeast Region presented a significant incidence peak in 2022 (1.73 cases/100,000 Inh). The Central-West region stands out for having a significantly lower incidence index than the other regions, and the maximum observed incidence was 0.52 cases/100,000 Inh in 2015 (Tab 3).

**Tab 3.** Leptospirosis incidence rate in Brazil and by region. The data detail the annual proportions (cases per 100,000 inhabitants) graphically represented in Figure 3.

| Year | Brazil | Central-West | North | Northeast | South | Southeast |
| --- | --- | --- | --- | --- | --- | --- |
| 2015 | 2.13 | 0.52 | 7.51 | 0.76 | 5.41 | 1.11 |
| 2016 | 1.49 | 0.47 | 2.71 | 0.58 | 4.14 | 1.13 |
| 2017 | 1.46 | 0.34 | 2.88 | 0.82 | 3.61 | 1.05 |
| 2018 | 1.48 | 0.44 | 2.72 | 0.83 | 3.44 | 1.16 |
| 2019 | 1.76 | 0.48 | 2.66 | 1.12 | 4.69 | 1.23 |
| 2020 | 0.87 | 0.35 | 1.64 | 0.48 | 1.76 | 0.75 |
| 2021 | 0.82 | 0.22 | 1.52 | 0.56 | 1.79 | 0.62 |
| 2022 | 1.49 | 0.31 | 1.67 | 1.73 | 2.60 | 1.14 |
| 2023 | 1.55 | 0.36 | 1.88 | 1.12 | 3.99 | 1.14 |
| 2024 | 1.88 | 0.47 | 1.88 | 1.12 | 6.53 | 1.05 |

**Fig 3.**
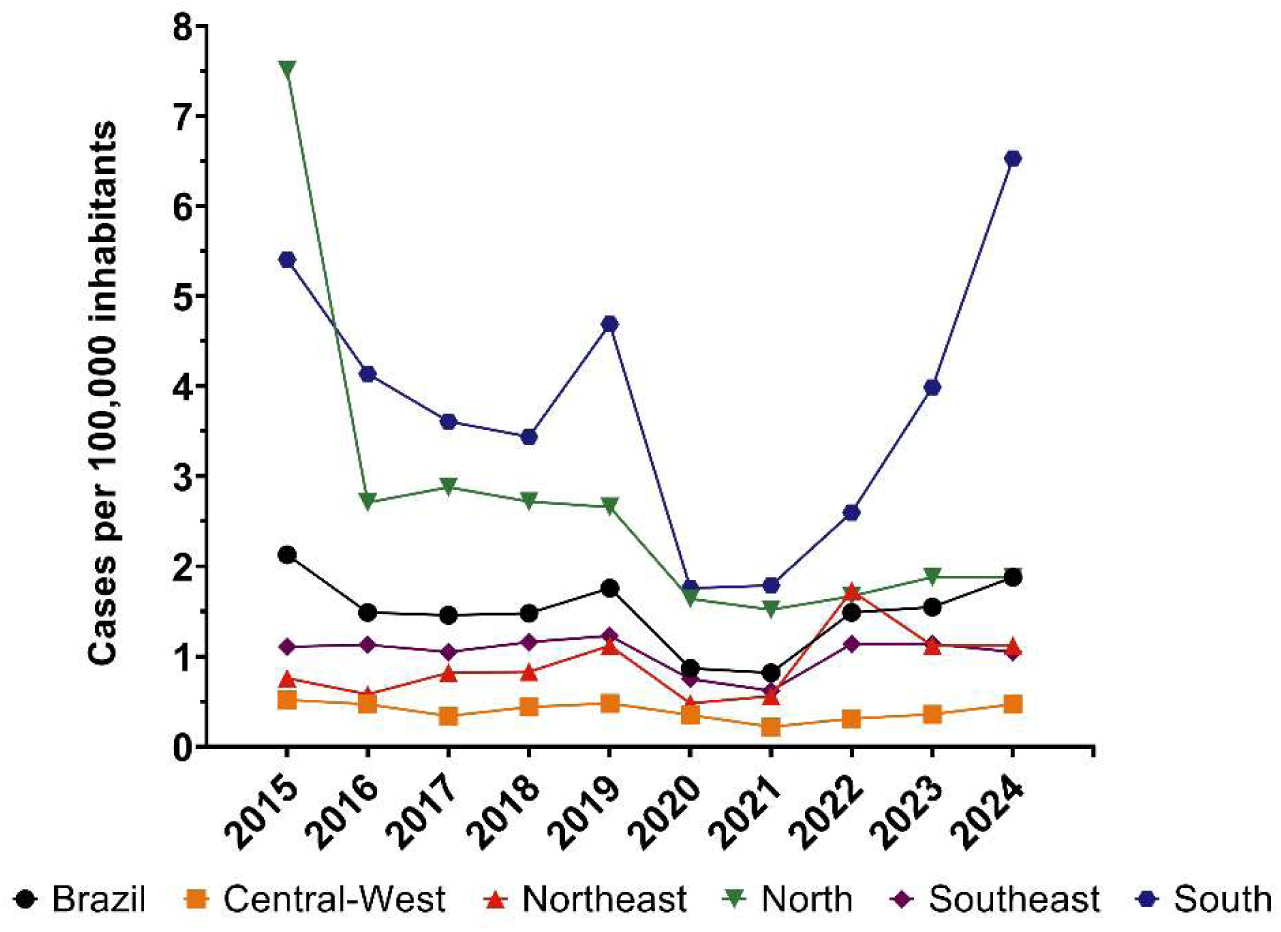
Leptospirosis incidence rate in Brazil and regions. The vertical axis indicates the annual proportion of cases per 100,000 inhabitants stratified by region and for the country (black line).

### Lethality index of leptospirosis in Brazil

The average lethality of leptospirosis in Brazil remained basically constant over the 10 years, varying from 7.79% to 10.58% of cases that evolved to death (Fig 4 and Tab 4). Interestingly the Northeast and Southeast regions had lethality indices above the national average, reaching 17.22 and 14.44, respectively. While the regions with the highest incidence of the disease, south and north, remained below the national average, with minimums of 4.11 in 2018 and 2.74 in 2015, respectively. The Central-West region presents a graphical large variation due to the low number of cases, which consequently is highly impacted by a low number of deaths.

**Tab 4.** Leptospirosis lethality rate in Brazil and by region.

| Leptospirosis Lethality in Brazilian Regions per Year |  |  |  |  |  |  |
| --- | --- | --- | --- | --- | --- | --- |
| Year | Brazil | Central-West | North | Northeast | South | Southeast |
| 2015 | 7.79 | 12.50 | 2.74 | 15.33 | 6.29 | 13.50 |
| 2016 | 8.91 | 10.81 | 5.21 | 16.36 | 5.27 | 12.64 |
| 2017 | 8.83 | 5.56 | 6.01 | 14.04 | 4.22 | 13.33 |
| 2018 | 9.42 | 20.00 | 7.27 | 14.04 | 4.11 | 12.93 |
| 2019 | 8.79 | 8.97 | 6.11 | 13.03 | 4.63 | 12.88 |
| 2020 | 10.58 | 5.26 | 6.84 | 17.22 | 5.08 | 14.44 |
| 2021 | 10.38 | 2.78 | 5.56 | 15.43 | 8.10 | 12.66 |
| 2022 | 10.02 | 1.92 | 7.84 | 12.90 | 7.15 | 10.53 |
| 2023 | 8.41 | 4.92 | 6.04 | 10.15 | 5.78 | 11.45 |
| 2024 | 8.88 | 6.17 | 6.81 | 12.69 | 5.93 | 13.50 |

The data detail the annual percentages of deaths graphically represented in Fig 4.

**Fig 4.**
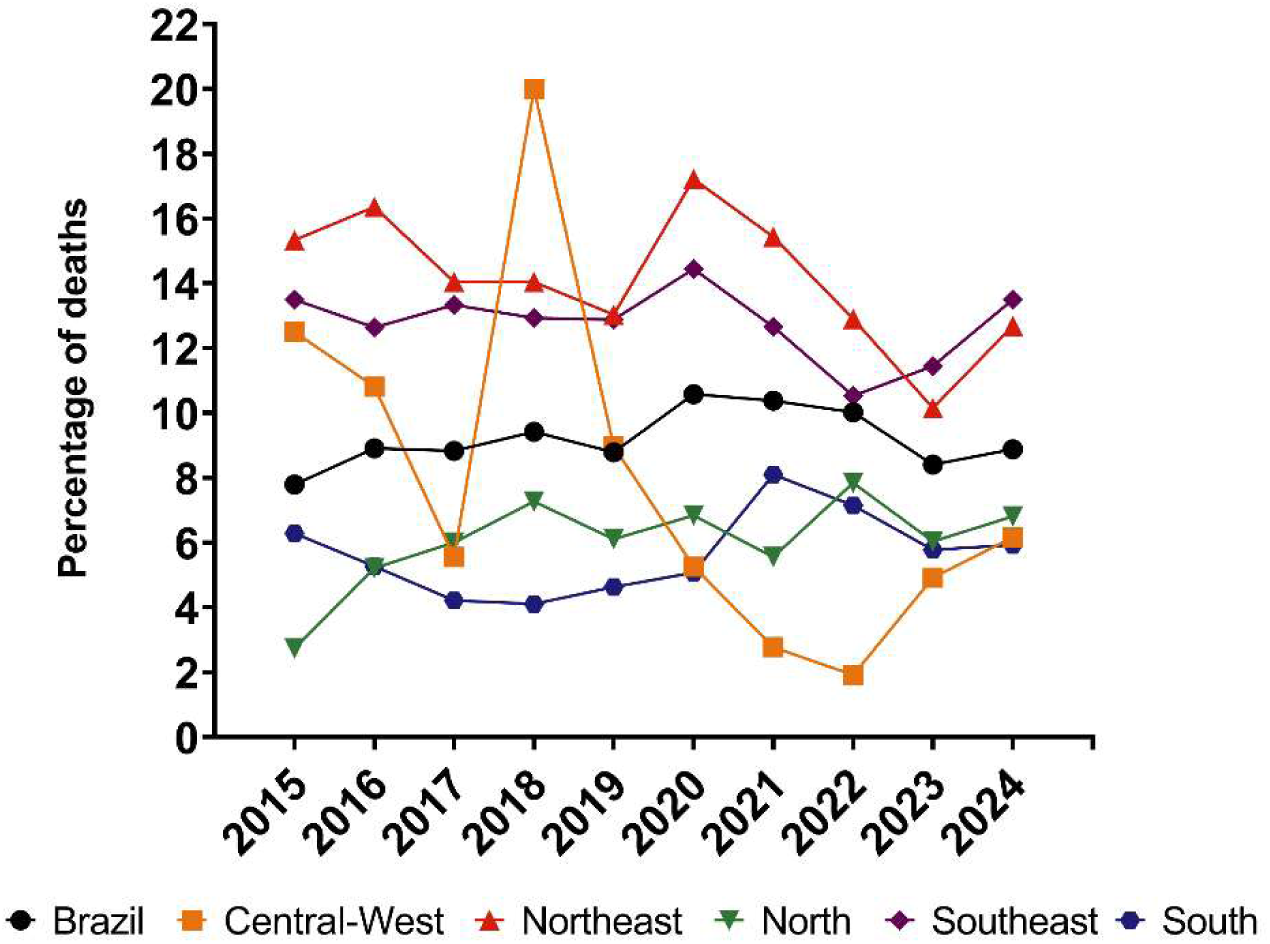
Leptospirosis lethality rate in Brazil and regions. The vertical axis shows the annual percentage of deaths relative to the total confirmed cases, stratified by region and for the national consolidated (black line).

### Distribution of leptospirosis cases according to environment and occupation

Among the 31,397 notified cases over the period, only 18,894 cases, corresponding to 60.1% of the total, reported the type of environment where the contamination occurred. Of the contaminations with indicated locations in Brazil, 64% occurred in residential environments, 26% in workplaces, and 10% during leisure activities (Fig 5 and Tab 5). The distribution pattern found in Brazil, is repeated across all regions. It is highlighted that in the north region, the number of domiciliary cases clearly exceeds the annual average, at 82%. Cases linked to the occupational environment are higher in the Central-West and South compared to the other regions, at 38% and 30%, respectively; however, it is important to consider that the Central-West has a small sample size, and therefore, the percentages may be unrepresentative. The cases related to leisure range from 13% in the Central-West and South regions to 3% in the North region.

**Tab 5.** Annual average of leptospirosis cases according to the probable environment of infection by region and federative unit in Brazil (2015–2024). The values represent the absolute average of the analyzed decade for notifications in domiciliary, work, and leisure environments.

| Subdivision | Domiciliary | Labor | Leisure |
| --- | --- | --- | --- |
| Brazil | 1212 | 494 | 183 |
| Central-West | 19 | 14 | 5 |
| Distrito Federal | 6 | 6 | 3 |
| Goiás | 6 | 4 | 2 |
| Mato Grosso | 4 | 5 | 2 |
| Mato Grosso do Sul | 3 | 1 | 2 |
| North | 287 | 55 | 8 |
| Acre | 153 | 25 | 2 |
| Amapá | 39 | 3 | 3 |
| Amazonas | 24 | 7 | 2 |
| Pará | 50 | 14 | 3 |
| Rondônia | 18 | 5 | 2 |
| Roraima | 2 | 2 | 1 |
| Tocantins | 4 | 2 | 3 |
| Northeast | 126 | 48 | 11 |
| Alagoas | 14 | 7 | 3 |
| Bahia | 23 | 10 | 2 |
| Ceará | 19 | 7 | 2 |
| Maranhão | 10 | 3 | 2 |
| Paraíba | 5 | 3 | 1 |
| Pernambuco | 42 | 11 | 3 |
| Piauí | 3 | 2 | 1 |
| Rio Grande do Norte | 4 | 4 | 2 |
| Sergipe | 10 | 6 | 2 |
| South | 451 | 243 | 103 |
| Paraná | 133 | 77 | 39 |
| Rio Grande do Sul | 196 | 97 | 38 |
| Santa Catarina | 123 | 69 | 27 |
| Southeast | 329 | 135 | 55 |
| Espírito Santo | 23 | 14 | 2 |
| Minas Gerais | 57 | 36 | 12 |
| Rio de Janeiro | 84 | 21 | 6 |
| São Paulo | 178 | 71 | 36 |

**Fig 5.**
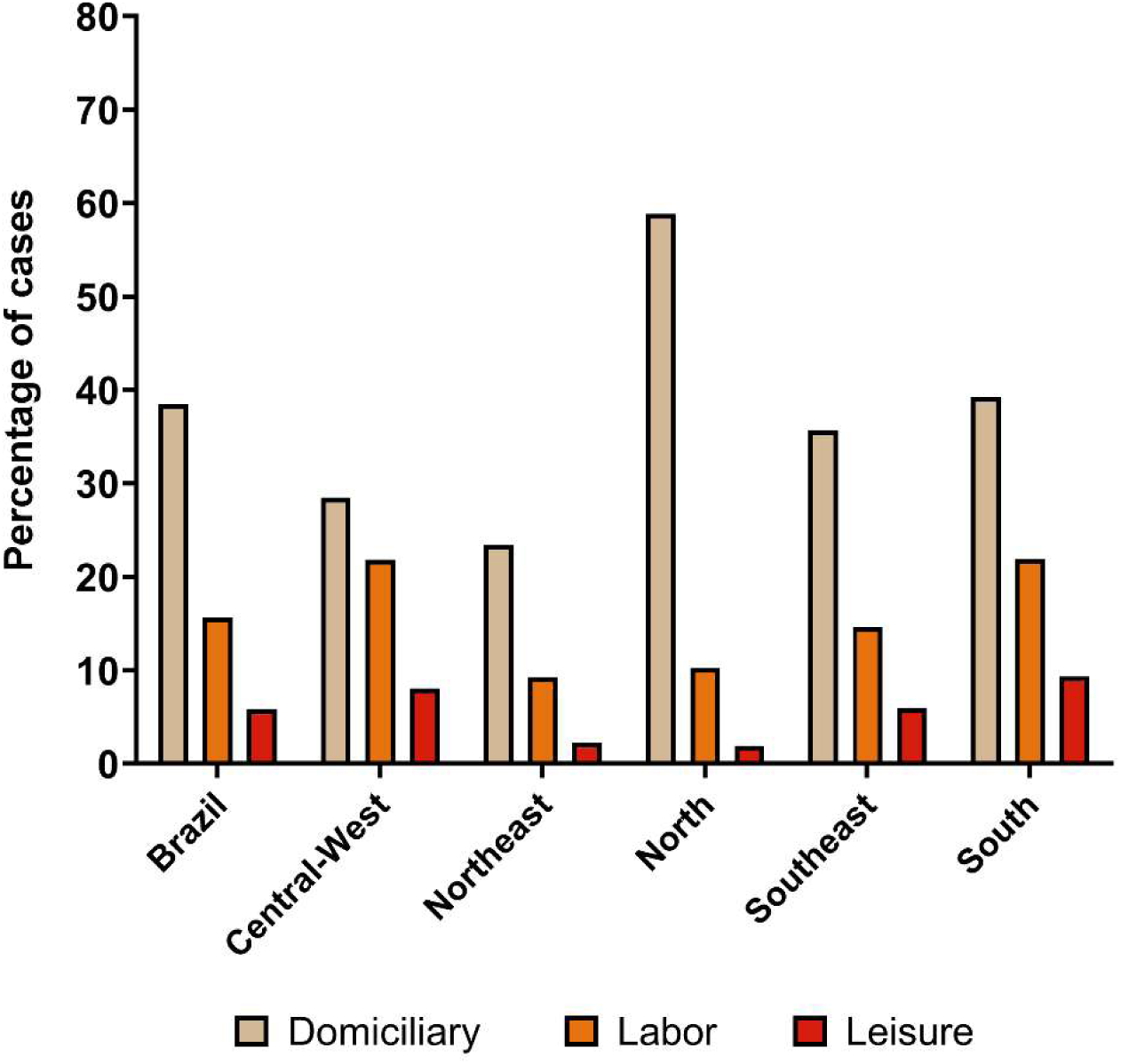
Proportion of leptospirosis cases according to the probable environment of infection in Brazil and regions (2015–2024). The vertical axis indicates the percentage of notifications in domiciliary, work, and leisure environments. The calculation exclusively considered the records with a valid location of infection, excluding blank or ignored data. The "other environments" category includes the percentage calculation; however, it was omitted from the graph due to the unspecificity of this variable in the registration forms.

According to the epidemiological surveillance panel, among the leptospirosis notifications that informed the occupation, it was identified that rural workers in agriculture, livestock farming, fishing, and other related activities accounted for 27.45% of the cases. Subsequently, jobs related to civil construction in general account for 18.63% of the cases. It is interesting to note that these two types of occupation, when added together, represent a significant portion of the cases with known occupation, accounting for 46.08%. Occupations related to industry, administrative, and health services account for the lowest percentage of cases, with about 3.92%. Furthermore, an expressive number of notified cases (16.67%) were not related to a specific occupation. It is important to emphasize that these percentages correspond only to the number of cases with the occupation declared in the notification and, furthermore, do not necessarily reflect a relationship with the location of contamination (Fig 6).

**Fig 6.**
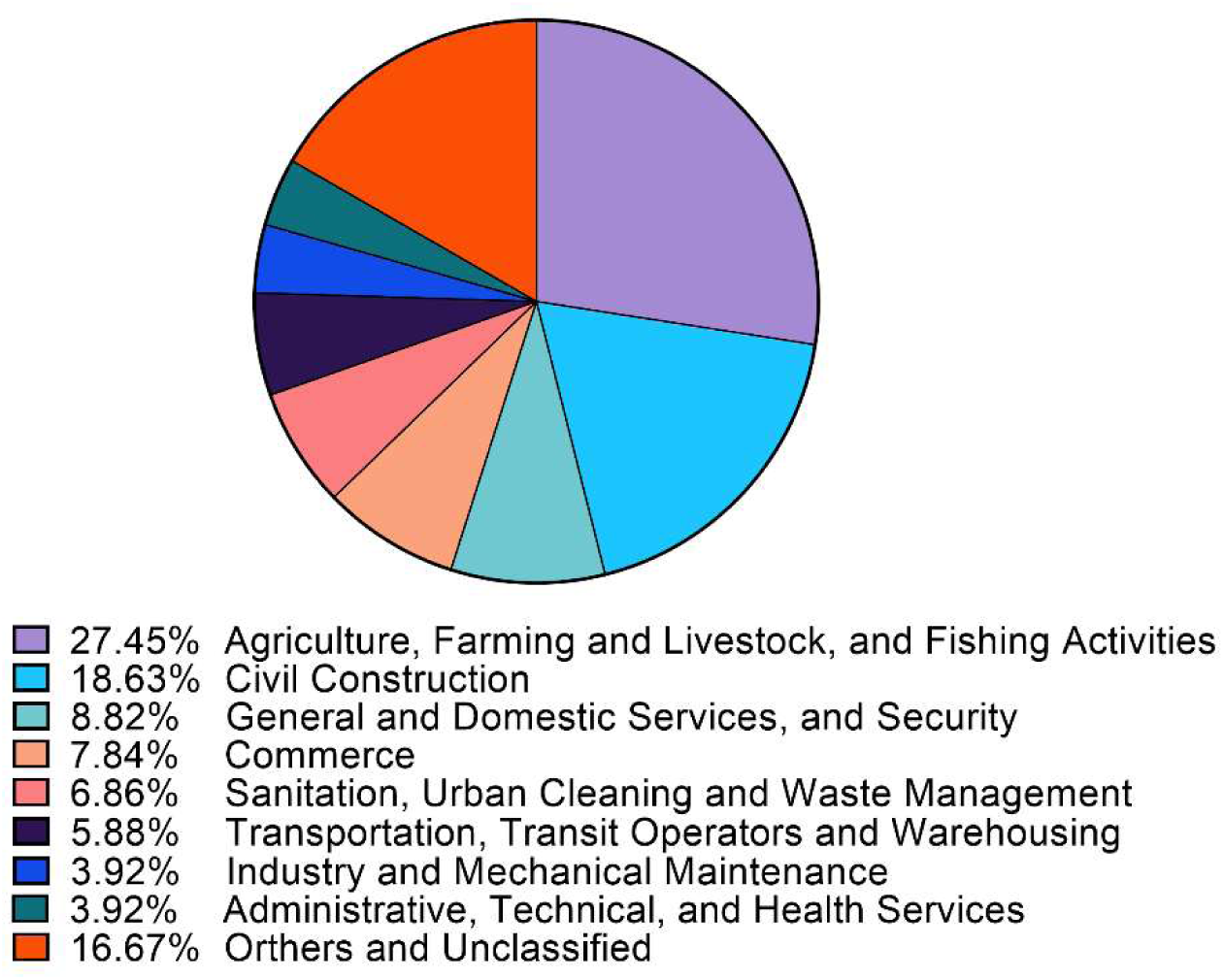
Percentage distribution of leptospirosis cases according to occupation (2015–2024). The illustrated categories represent the grouping of the 100 most frequent occupations registered in the Epidemiological Surveillance Panel, corresponding to 83.33% of the total notifications in the period. The "Others and unclassified" category (16.67%) encompasses the residual volume of occupations not listed in the main ranking or not specified.

### Association between leptospirosis cases and education level

To assess the influence of education level on leptospirosis incidence, it was used the International Standard Classification of Education (ISCED). The illiterate Brazilian population, classified as ISCED 0, together with the population with low education level, ISCED 1-2, presented incidence rates of 1.31 and 1.53 cases per 100,000 inhabitants, respectively. For the population with medium and high education levels, ISCED 3-4 and 5-6, rates of 0.87 and 0.39 cases per 100,000 inhabitants were calculated, respectively. This same pattern of reduction in incidence with the increase in education is observed in different proportions among the Brazilian regions and states, being notably evidenced in the south region, which has an incidence rate of 3.92 and 4.5 per 100,000 inhabitants for categories 0 and 1-2, while for categories 3-4 a reduction to 2.68 per 100,000 inhabitants was observed, which becomes even more notable in categories 5-6 with 1.08 cases per 100,000 inhabitants (Fig 7 and Tab 6).

**Tab 6.**
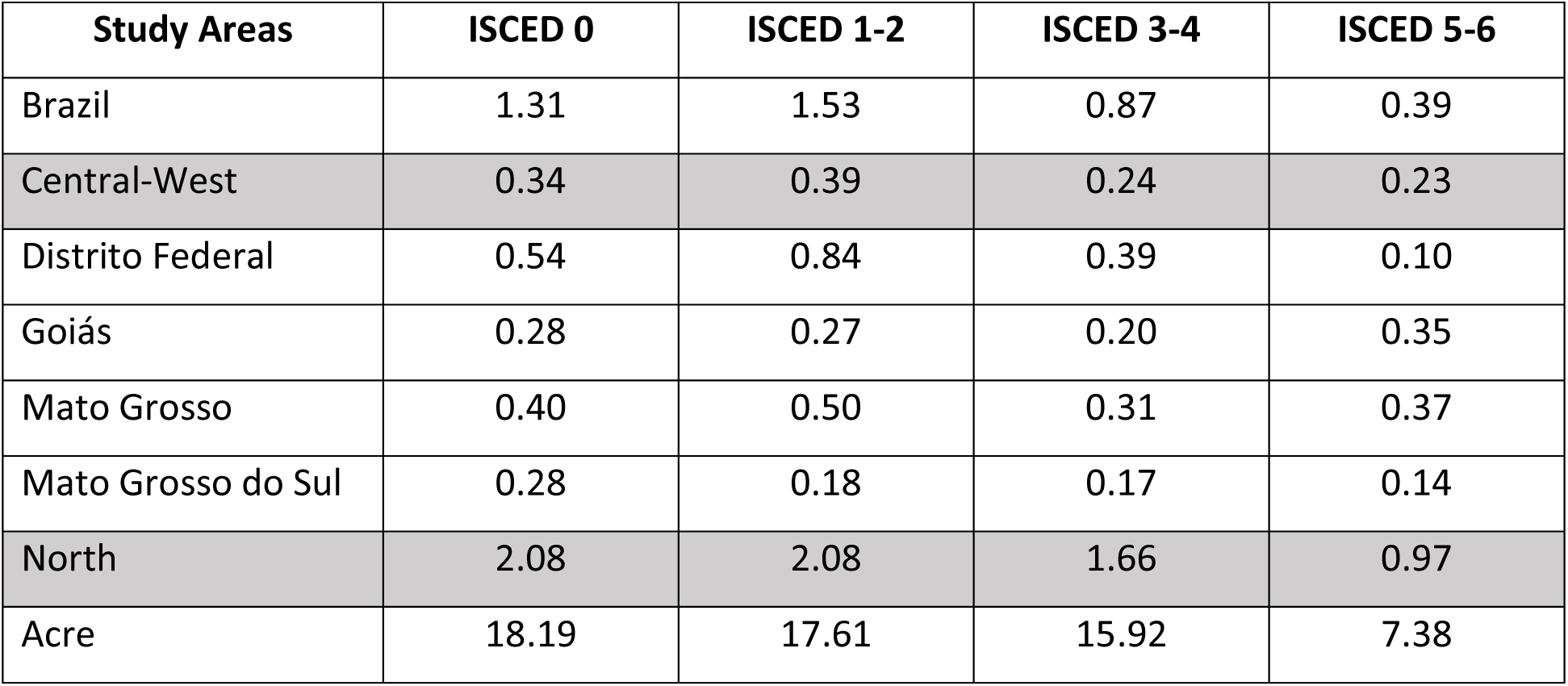
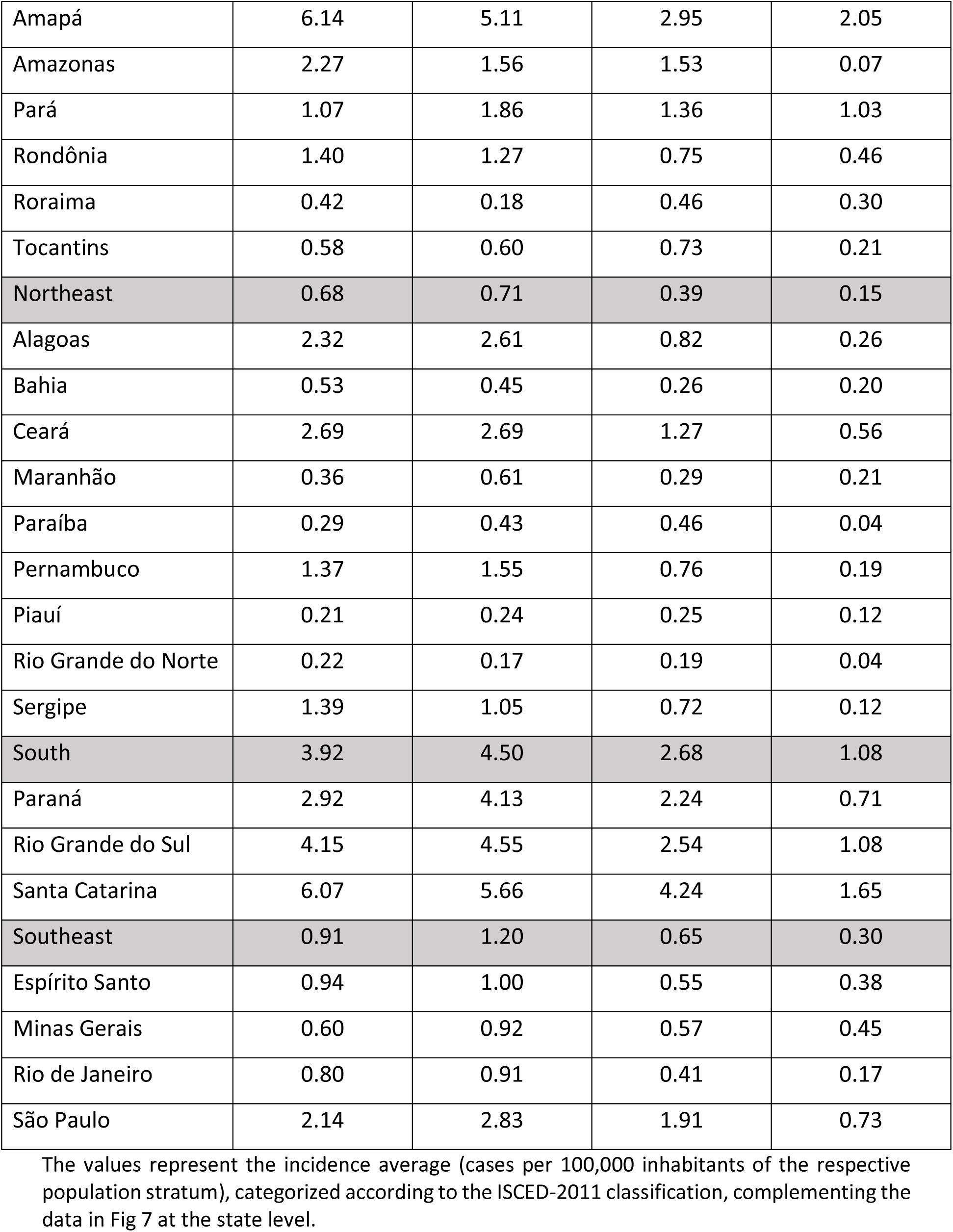
Leptospirosis incidence rate according to the educational level by region and federative unit in Brazil (2015–2024).

| Study Areas | ISCED 0 | ISCED 1-2 | ISCED 3-4 | ISCED 5-6 |
| --- | --- | --- | --- | --- |
| Brazil | 1.31 | 1.53 | 0.87 | 0.39 |
| Central-West | 0.34 | 0.39 | 0.24 | 0.23 |
| Distrito Federal | 0.54 | 0.84 | 0.39 | 0.10 |
| Goiás | 0.28 | 0.27 | 0.20 | 0.35 |
| Mato Grosso | 0.40 | 0.50 | 0.31 | 0.37 |
| Mato Grosso do Sul | 0.28 | 0.18 | 0.17 | 0.14 |
| North | 2.08 | 2.08 | 1.66 | 0.97 |
| Acre | 18.19 | 17.61 | 15.92 | 7.38 |
| Amapá | 6.14 | 5.11 | 2.95 | 2.05 |
| Amazonas | 2.27 | 1.56 | 1.53 | 0.07 |
| Pará | 1.07 | 1.86 | 1.36 | 1.03 |
| Rondônia | 1.40 | 1.27 | 0.75 | 0.46 |
| Roraima | 0.42 | 0.18 | 0.46 | 0.30 |
| Tocantins | 0.58 | 0.60 | 0.73 | 0.21 |
| Northeast | 0.68 | 0.71 | 0.39 | 0.15 |
| Alagoas | 2.32 | 2.61 | 0.82 | 0.26 |
| Bahia | 0.53 | 0.45 | 0.26 | 0.20 |
| Ceará | 2.69 | 2.69 | 1.27 | 0.56 |
| Maranhão | 0.36 | 0.61 | 0.29 | 0.21 |
| Paraíba | 0.29 | 0.43 | 0.46 | 0.04 |
| Pernambuco | 1.37 | 1.55 | 0.76 | 0.19 |
| Piauí | 0.21 | 0.24 | 0.25 | 0.12 |
| Rio Grande do Norte | 0.22 | 0.17 | 0.19 | 0.04 |
| Sergipe | 1.39 | 1.05 | 0.72 | 0.12 |
| South | 3.92 | 4.50 | 2.68 | 1.08 |
| Paraná | 2.92 | 4.13 | 2.24 | 0.71 |
| Rio Grande do Sul | 4.15 | 4.55 | 2.54 | 1.08 |
| Santa Catarina | 6.07 | 5.66 | 4.24 | 1.65 |
| Southeast | 0.91 | 1.20 | 0.65 | 0.30 |
| Espírito Santo | 0.94 | 1.00 | 0.55 | 0.38 |
| Minas Gerais | 0.60 | 0.92 | 0.57 | 0.45 |
| Rio de Janeiro | 0.80 | 0.91 | 0.41 | 0.17 |
| São Paulo | 2.14 | 2.83 | 1.91 | 0.73 |

**Fig 7.**
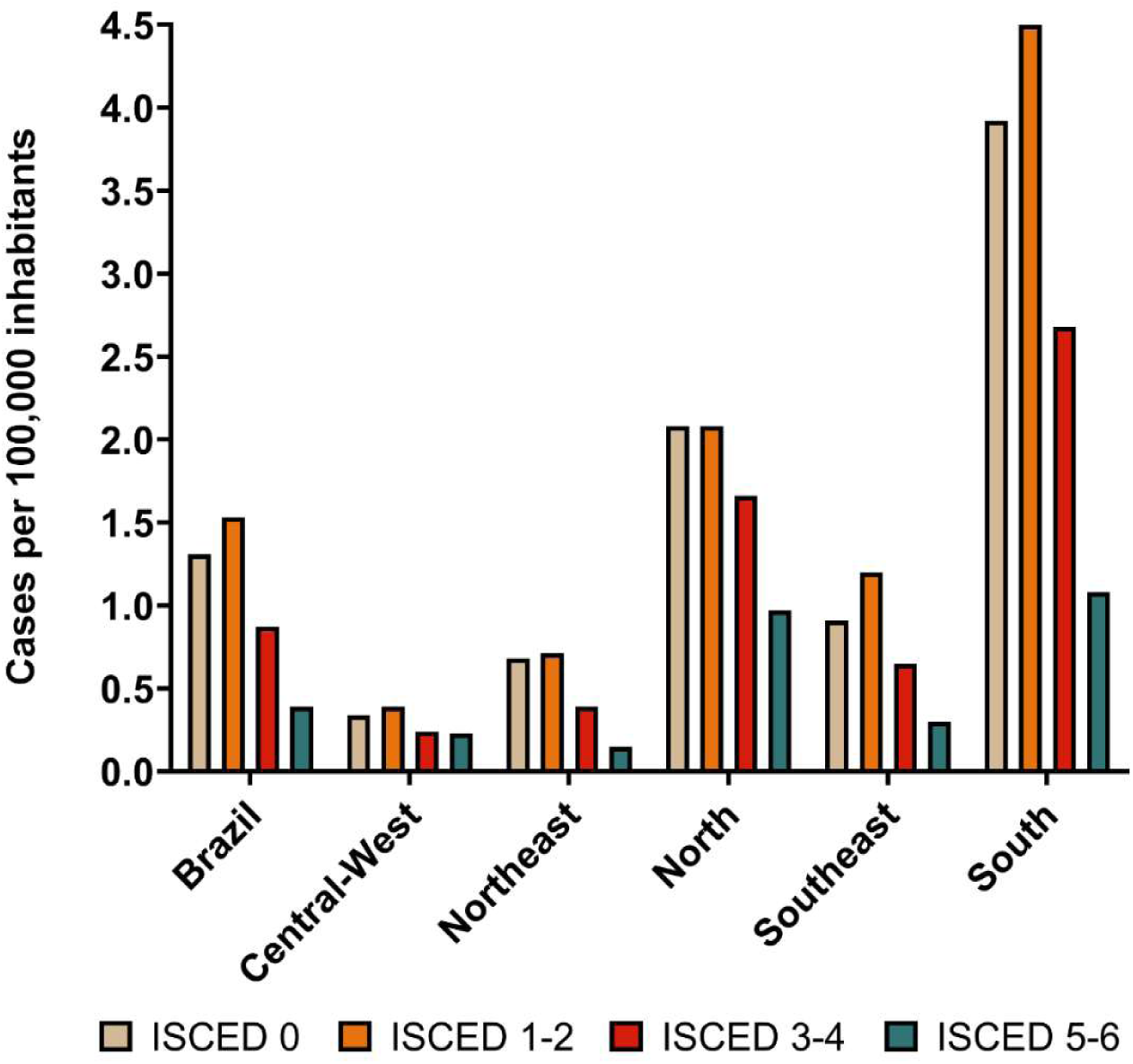
Leptospirosis incidence rate according to educational level in Brazil and regions (2015–2024). The vertical axis shows the average incidence (cases per 100,000 inhabitants), stratified by each educational group’s population size. The categories were standardized according to the International Standard Classification of Education (ISCED-2011): ISCED 0 (no formal education/illiteracy); ISCED 1-2 (elementary and incomplete high school); ISCED 3-4 (complete high school and incomplete higher education); and ISCED 5-6 (complete technical level or higher education).

### Climatic influence on the distribution of leptospirosis cases

The analysis of the spatiotemporal correlation between the leptospirosis incidence rate and the climatic anomalies of the El Niño-Southern Oscillation (ENSO), evaluated from the Oceanic Niño Index (ONI/NOAA) data, revealed a heterogeneous response pattern across the Brazilian territory (Fig 8). A positive, statistically significant correlation was observed in most of the states of the South and Southeast regions. This finding indicates that the intensification of El Niño acts as a positive modulator of the disease in these areas, associated with a tendency to increase the number of cases. In contrast, some states in the Northeast region, notably Ceará, Rio Grande do Norte, Paraíba, and Sergipe, demonstrated a negative correlation, which may indicate a decrease in incidence during periods of heightened climatic phenomenon intensity

**Fig 8.**
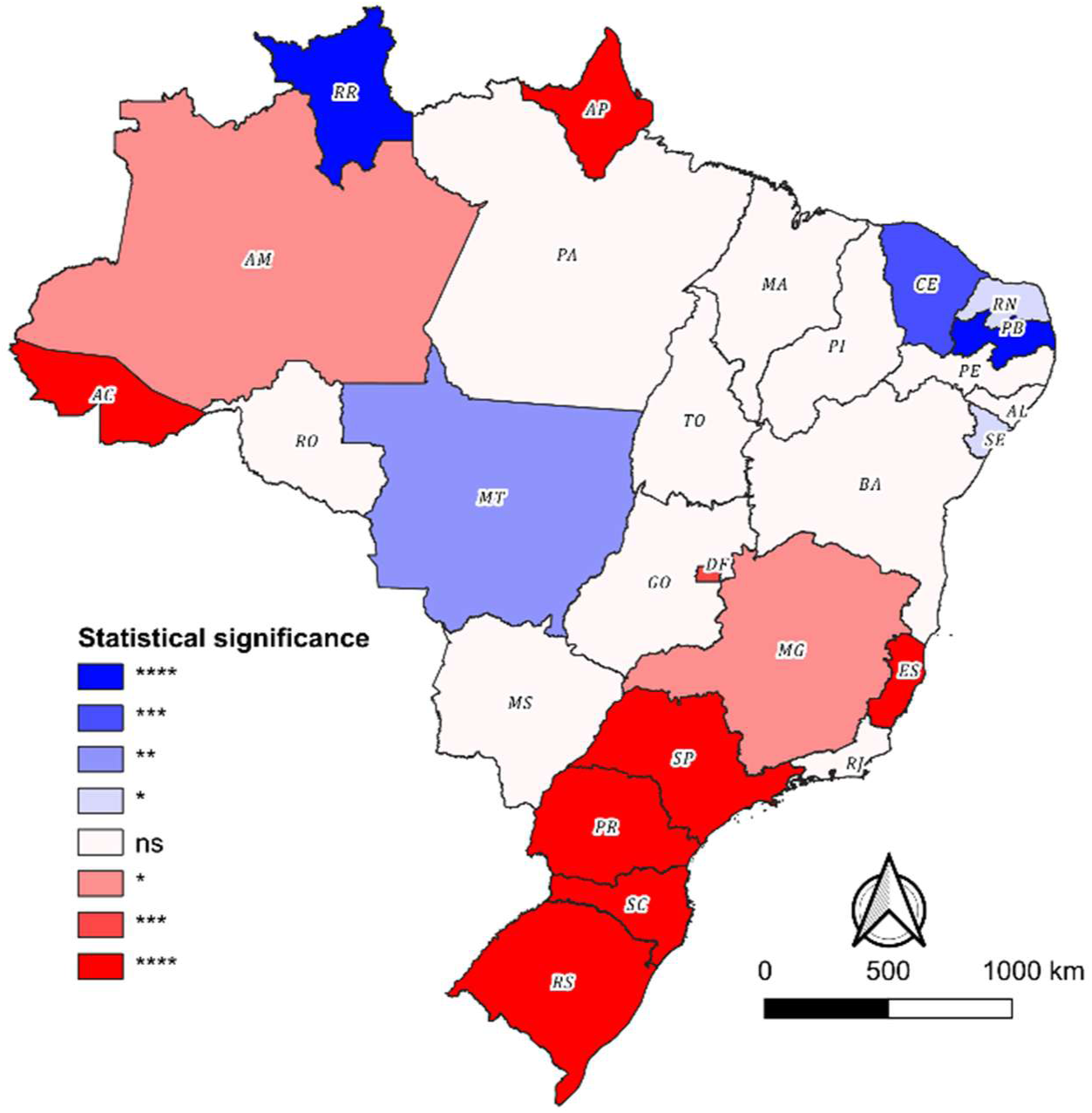
Spatial correlation between the ENSO climatic phenomenon and leptospirosis incidence in Brazil (2015–2024). The choropleth map illustrates Spearman’s correlation coefficients between the El Niño oceanic index and state-level incidence rates. States in red indicate a positive correlation (an increase in incidence associated with the intensification of the phenomenon), whereas those in blue indicate a negative correlation. Color saturation and markings denote the statistical significance of the two-tailed tests (ns, non-significant; *, p < 0.05; **, p < 0.01; ***, p < 0.001; ****, p < 0.0001).

It was also verified that several federative units did not present statistical significance for this analysis or exhibited divergent intraregional patterns. In the North region, for example, the state of Roraima demonstrated a strong negative association, while Acre and Amapá exhibited a robust positive correlation. The absence of a uniform pattern and statistical significance in a large part of the territory reinforces the multifactorial etiology of leptospirosis. The incidence of the disease is not exclusively determined by macroclimatic variability and precipitation regimes associated with ENSO; in regions where there is no significant correlation, it is probable that local socioenvironmental and infrastructural determinants play a preponderant role in the seasonality of the infection.

The analysis of the historical series of the geographic distribution of leptospirosis incidence in Brazil corroborates the complexity of this scenario (Fig 9). A persistently high disease burden is observed in states such as Acre and Amapá, which characterizes leptospirosis as highly endemic in these locations. The South region, in turn, exhibits consistently high incidence rates with periods of exacerbation that constitute epidemic outbreaks. Additionally, states such as Pernambuco and Espírito Santo present a pattern of occurrence of lesser magnitude, marked by sporadic epidemic episodes.

**Fig 9.**
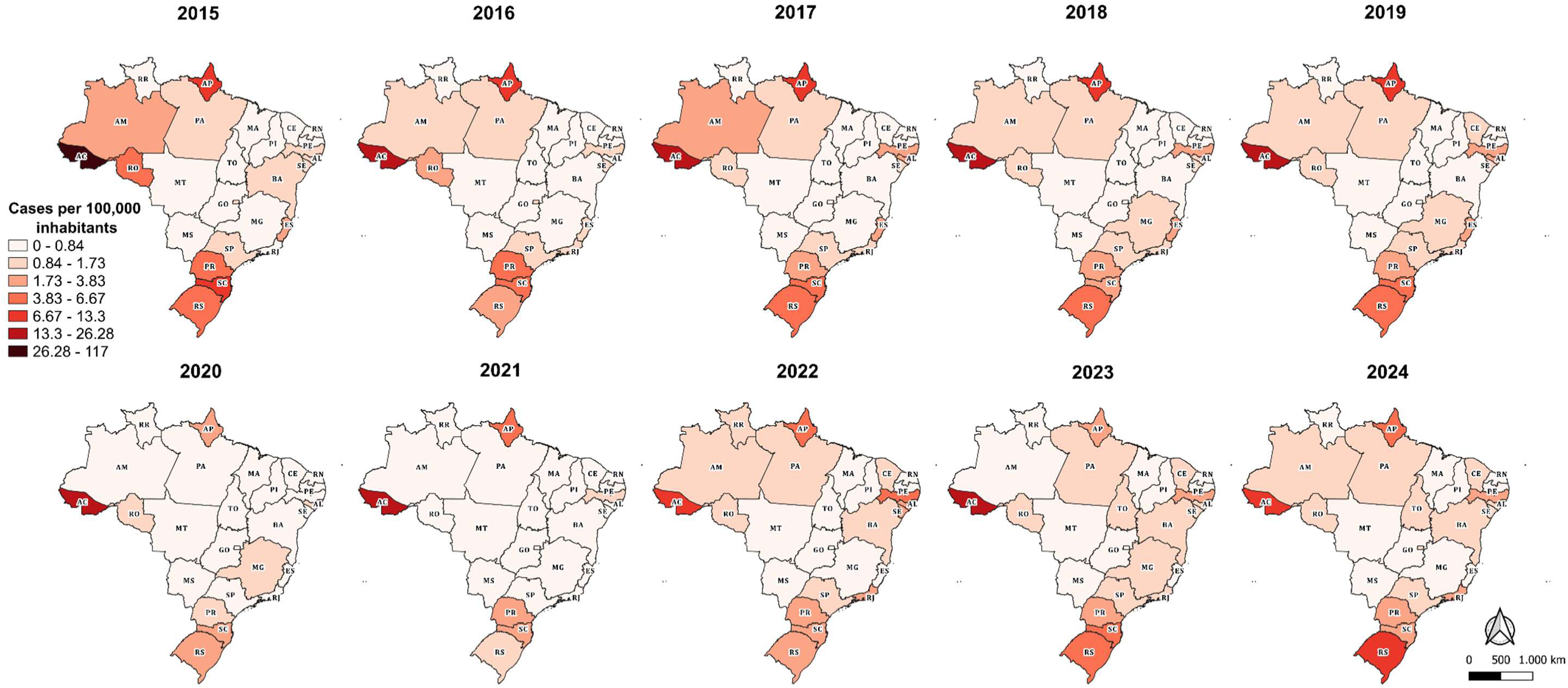
Spatiotemporal distribution of leptospirosis incidence in Brazil by federative unit (2015–2024). The series of annual choropleth maps illustrates the incidence rate (cases per 100,000 inhabitants). To ensure visual comparability throughout the decade and to attenuate the visual flattening caused by high-incidence outliers (e.g., the state of Acre), the continuous color scale was stratified into six classes using the recursive median division method applied to the total dataset for the period. Darker shades of red indicate the highest registered incidence rates.

## DISCUSSION

Although leptospirosis is internationally recognized as a neglected tropical disease, in Brazil, despite being considered an important public health problem, it was not included in the epidemiological bulletin of neglected diseases (16). Therefore, there is no Brazilian funding program aimed at the intervention of this disease. However, taking into account the potential years of life lost and the hospital costs of leptospirosis in Brazil (17), this disease deserves prominence. In this study, a total of 31,397 leptospirosis cases were identified over the last 10 years, with an annual average of 3,140 cases. The annual average of cases apparently did not show large variations over long periods, since a previous analysis conducted from 2000 to 2015 (18), reported an average of 3,810 annual cases. This average coincided with the data analyzed from 2007 to 2017, with 3,846 cases (19). Furthermore, considering the average annual number of cases observed from 1996 to 2005 (3,165 notifications), we can suggest that the number of cases in Brazil remained stable over these years (20). Although this maintenance is already expected due to the absence of a vaccine for human use (21) and the lack of significant improvements in the sanitary conditions of the cities, we must be aware that the lack of diagnosis in the initial phase of the disease can lead to underreporting of data. The study conducted from 1996 to 2005 showed that Brazil was responsible for about 86% of the cases across South America (20), emphasizing the importance of this zoonosis to the country.

An interesting observation in the temporal dynamics of this survey was a reduction in notifications across all regions during the COVID-19 pandemic. This pattern was not exclusive to leptospirosis, with a decrease observed in other compulsorily notifiable diseases in Brazil (22, 23) and in other countries, such as China and Australia (24, 25). This strongly suggests that, due to the health system’s focus on the pandemic, there was a failure in notifications, a premise reinforced by the maintenance of the lethality index during the period, particularly in the Southeast and Northeast.

The evaluation of the epidemiological determinants of leptospirosis requires a critical reflection on the effect of geographic scale. While the present analysis at the national level offers a comprehensive panorama, studies at the municipal scale tend to provide greater precision in capturing socioenvironmental nuances, as demonstrated in a previous study in Rio de Janeiro (26). However, extrapolating hyper-localized analyses poses limitations given Brazil’s continental dimensions. In this sense, the formulation of a regional and systemic vision is indispensable.

The analysis of the location of contamination reveals that the domiciliary environment represents the main route of exposure (64%), followed by the occupational environment (26%) and recreational activities (10%). This profile reflects the dynamics of low- and middle-income countries, where precarious sanitary infrastructure is a determinant of the infection (27, 28), contrasting with high-income nations, where sports and leisure predominate. In Brazilian urban settings, proximity to open sewers and the accumulation of garbage promote the proliferation of synanthropic rodents (*Rattus norvegicus*), thereby consolidating the domiciliary as the epicenter of transmission among marginalized populations (29, 30).

Regionally, the disparity is evident in the North, where the residential environment accounts for 82% of contaminations, indicating a critical deficit in sanitation (31). On the other hand, the Central-West and South had higher proportions of occupational cases (38% and 30%), indicating an interface with agribusiness. From an occupational perspective, the predominance of rural (27.45%) and civil construction workers (18.63%) underlines the overlap between unsanitary work and environmental risk (32). This vulnerability is corroborated by education level (ISCED): the highest incidence occurs in illiterate populations (1.31/100,000) and in those with low education (1.53/100,000), consolidating the premise that education and income determine socio-spatial resilience (27, 33). The drastic decline of cases in the most educated strata evidences that favored populations inhabit zones with urban infrastructure that acts as a barrier against floods (34, 35). Furthermore, higher education commonly generates formal employment with higher remuneration; it was calculated that for every one-dollar increase in daily per capita expenditure per domiciliary, the chances of primary infection are reduced by 50% (30).

Regarding clinical outcomes, no direct relationship between incidence rate and lethality was observed across regions. In general, the North and South stand out for presenting high incidence and low lethality. In contrast, the largest number of cases in urban areas occurred in the Northeast (18), which had the lowest incidence rate but the highest lethality rate during the period. This scenario suggests a specific failure in the early diagnosis of the disease. The state of Sergipe draws attention due to the elevated number of deaths: A study conducted in the state pointed out that, of 516 notified cases (2007-2022), 140 evolved to death (lethality of 27.2%) (36). Similar to previous observations (37, 38), the highest prevalence occurs in men of economically active age. The interregional lethality discrepancy raises the hypothesis that areas with higher mortality, such as the Southeast, may have a greater circulation of highly virulent strains, such as those of the Icterohaemorrhagiae serogroup; however, the scarcity of regionalized genomic surveillance data limits the definitive confirmation of these dynamics.

The high lethality in the Northeast also reflects the interaction between inadequate infrastructure and climatic factors (39). Globally, leptospirosis outbreaks are closely associated with extreme weather events (40). In this context, the analysis of the spatiotemporal correlation between leptospirosis incidence and ENSO anomalies reveals modulation by Brazil’s macroclimate (41, 42). In the South and Southeast regions, the significant positive correlation corresponds to atmospheric blocking associated with El Niño, which generates severe rainfall anomalies, thereby elevating the circulation of *Leptospira* sp. This mechanism corroborates studies that demonstrated a 31.5% increase in hospitalizations in São Paulo for every 20 mm of rain (43). In the South, these events induce the formation of risk clusters of leptospirosis dependent on hydrological disasters (44, 45), culminating in catastrophic outbreaks, such as the one in Rio Grande do Sul in 2024 (46, 47). In contrast, the negative correlation in the Northeast occurs because El Niño induces strong atmospheric subsidence (a descending air flow that suppresses cloud formation), resulting in severe droughts. These arid conditions reduce the contact between reservoir hosts and accidental human hosts, thereby lowering the incidence.

The Amazon basin exhibits a complex epidemiological mosaic. The state of Acre draws attention due to the alarming number of persistent cases. The positive climatic correlation reflects the dynamics of the Acre River, with recurrent overflow episodes. Associated factors, such as disorganized urban development along the riverbanks and increased reservoir density, including rodents and capybaras (48), favor local transmission. In 2015, Acre faced the largest flood in its history (49), which justified the extreme peak of notifications. Interestingly, in the second-worst overflow, in 2024, the average of notifications remained similar to previous years. This epidemiological plateau, allied with the state’s history of declaring environmental emergencies (50), suggests the effect of continuous, more active epidemiological surveillance in a known endemic region, whose peaks characteristically occur in March (51). The severity of these floods demands continuous strategies, such as the Emergency Plan for Coping with Floods (52). In parallel, Amapá also stands out due to the high endemism in the Guianas region, which actually consisted of underreporting and clinical confounding with malaria and arboviruses (53).

This study presents inherent limitations arising from its retrospective nature and reliance on secondary data from SINAN. Furthermore, the accuracy of population-based epidemiological analyses is strictly conditioned by the quality and completeness of compulsory notification forms, which are frequently affected by a high proportion of missing or blank fields (54). Added to this are the regional disparities in laboratory infrastructure and in the continuous training of health surveillance teams. Such asymmetries can introduce underreporting biases and distort the real magnitude of the incidence, evidencing the importance of professional qualification and strengthened notification systems to support more precise public health policies.

The dynamics of leptospirosis in Brazil transcend its infectious etiology, consolidating itself as an acute marker of socioenvironmental vulnerability and infrastructural inequity. The predominance of infections in the domiciliary environment, combined with higher incidence among populations with low levels of education and in occupational sectors historically exposed to unsanitary conditions, evidences the urgency of public policies aimed at universalizing basic sanitation. Additionally, the strong modulation of the ENSO on the spatiotemporal distribution of cases demonstrates that the epidemic risk is geographically heterogeneous and closely linked to climatic extremes. To effectively mitigate the burden of this zoonosis, it is imperative that the Brazilian public health system adopts an intersectoral approach. This requires not only the continuous strengthening of epidemiological surveillance and diagnostic capacity to combat underreporting, but also the integration of climate forecasting models to formulate regionalized early warning systems that anticipate and mitigate outbreaks in areas of chronic hydrological risk.

## Data Availability

All relevant data are within the manuscript and its Supporting Information files.

## Acknowledgements

Supported by FAPESP (Fundação de Amparo à Pesquisa do Estado de São Paulo – “Foundation for Research Support of the State of São Paulo”), CNPq (Conselho Nacional de Desenvolvimento Científico e Tecnológico - "National Counsel of Technological and Scientific Development"). CAPES (Coordenação de Aperfeiçoamento de Pessoal de Nível Superior - Coordination for the Improvement of Higher Education Personnel), and Fundação Butantan (Butantan Foundation).

## Conflicts of Interest

The authors declare no conflicts of interest. The funders had no role in the design of the study; in the collection, analyses, or interpretation of data; in the writing of the manuscript; or in the decision to publish the results.

## Notes

### Competing Interest Statement

The authors have declared no competing interest.

### Funding Statement

The author(s) received no specific funding for this work.

### Author Declarations

it is a retrospective study based on secondary data in the public domain, strictly anonymous and without the possibility of individual identification, the present work is exempt from review by a Research Ethics Committee, in accordance with the guidelines of the National Health Council (CNS) and the Brazilian General Data Protection Law (LGPD – Law No. 13,709/2018).

## REFERENCES

1. Soo ZMP, Khan NA, Siddiqui R. Leptospirosis: Increasing importance in developing countries. Acta Trop. 2020;201:105183.

2. Cunha M, Costa F, Ribeiro GS, Carvalho MS, Reis RB, Nery N, et al. Rainfall and other meteorological factors as drivers of urban transmission of leptospirosis. PLoS Negl Trop Dis. 2022;16(4):e0007507.

3. Vincent AT, Schiettekatte O, Goarant C, Neela VK, Bernet E, Thibeaux R, et al. Revisiting the taxonomy and evolution of pathogenicity of the genus Leptospira through the prism of genomics. PLoS Negl Trop Dis. 2019;13(5):e0007270.

4. Bharti AR, Nally JE, Ricaldi JN, Matthias MA, Diaz MM, Lovett MA, et al. Leptospirosis: a zoonotic disease of global importance. The Lancet infectious diseases. 2003;3(12):757–71.

5. Adler B, de la Peña Moctezuma A. Leptospira and leptospirosis. Vet Microbiol. 2010;140(3-4):287–96.

6. Picardeau M, Brenot A, Saint Girons I. First evidence for gene replacement in Leptospira spp. Inactivation of L. biflexa flaB results in non-motile mutants deficient in endoflagella. Mol Microbiol. 2001;40(1):189–99.

7. Cullen PA, Haake DA, Adler B. Outer membrane proteins of pathogenic spirochetes. FEMS Microbiol Rev. 2004;28(3):291–318.

8. Levett PN. Leptospirosis. Clin Microbiol Rev. 2001;14(2):296–326.

9. Haake DA, Levett PN. Leptospirosis in humans. Curr Top Microbiol Immunol. 2015;387:65–97.

10. Plank R, Dean D. Overview of the epidemiology, microbiology, and pathogenesis of Leptospira spp. in humans. Microbes and infection. 2000;2(10):1265–76.

11. Faine S, Adler B, Bolin C, Perolat P. Leptospira and leptospirosis, Melbourne. Australia: MediSci. 1999;259.

12. Laurichesse H, Gourdon F, Smits HL, Abdoe TH, Estavoyer JM, Rebika H, et al. Safety and immunogenicity of subcutaneous or intramuscular administration of a monovalent inactivated vaccine against Leptospira interrogans serogroup Icterohaemorrhagiae in healthy volunteers. Clin Microbiol Infect. 2007;13(4):395–403.

13. Yanagihara Y, Villanueva SY, Yoshida S, Okamoto Y, Masuzawa T. Current status of leptospirosis in Japan and Philippines. Comp Immunol Microbiol Infect Dis. 2007;30(5-6):399–413.

14. Martínez R, Pérez A, Quiñones MeC, Cruz R, Alvarez A, Armesto M, et al. [Efficacy and safety of a vaccine against human leptospirosis in Cuba]. Rev Panam Salud Publica. 2004;15(4):249–55.

15. OECD E, UNESCO Institute, for Statistics. ISCED 2011 Operational Manual: Guidelines for Classifying National Education Programmes and Related Qualifications. Paris: OECD Publishing; 2015.

16. Brasil MdS. Doenças Tropicais Negligenciadas no Brasil: morbimortalidade e resposta nacional no contexto dos Objetivos de Desenvolvimento Sustentável 2016-2020. In: Ambiente SdVeSe, editor. Brasília: Ministério da Saúde; 2024.

17. Souza VM, Arsky MeL, Castro AP, Araujo WN. Years of potential life lost and hospitalization costs associated with leptospirosis in Brazil. Rev Saude Publica. 2011;45(6):1001–8.

18. Galan DI, Roess AA, Pereira SVC, Schneider MC. Epidemiology of human leptospirosis in urban and rural areas of Brazil, 2000-2015. PLoS One. 2021;16(3):e0247763.

19. Nardoni Marteli A, Guasselli LA, Diament D, Wink GO, Vasconcelos VV. Spatio-temporal analysis of leptospirosis in Brazil and its relationship with flooding. Geospat Health. 2022;17(2).

20. Costa F, Martinez-Silveira MS, Hagan JE, Hartskeerl RA, Dos Reis MG, Ko AI. Surveillance for leptospirosis in the Americas, 1996-2005: a review of data from ministries of health. Rev Panam Salud Publica. 2012;32(3):169–77.

21. Barazzone GC, Teixeira AF, Azevedo BOP, Damiano DK, Oliveira MP, Nascimento ALTO, et al. Revisiting the Development of Vaccines Against Pathogenic. Front Immunol. 2021;12:760291.

22. de Souza Matsumura ES, Carneiro TX, Feio ECG, Guedes JA, Xavier MB. Impacto da pandemia COVID-19 nas notificações das doenças infecciosas no município de Belém–PA. Revista Eletrônica Acervo Saúde| ISSN.2178:2091.

23. Borges PKdO, Martins CM, Stocco C, Zuber JFS, Borges WS, Muller EV, et al. Impacto da COVID-19 sobre doenças de notificação compulsória: um estudo de série temporal. Revista da Escola de Enfermagem da USP. 2024;58:e20240098.

24. Sohail A, Cheng AC, McGuinness SL, Leder K. The epidemiology of notifiable diseases in Australia and the impact of the COVID-19 pandemic, 2012-2022. BMC Glob Public Health. 2024;2(1):1.

25. Yu Y, Shen L. Changes in the incidence of notifiable infectious diseases before, during, and after the COVID-19 pandemic in Mainland China. BMC Infect Dis. 2025;26(1):36.

26. Gracie R, Barcellos C, Magalhães M, Souza-Santos R, Barrocas PR. Geographical scale effects on the analysis of leptospirosis determinants. Int J Environ Res Public Health. 2014;11(10):10366–83.

27. Reis RB, Ribeiro GS, Felzemburgh RD, Santana FS, Mohr S, Melendez AX, et al. Impact of environment and social gradient on Leptospira infection in urban slums. PLoS Negl Trop Dis. 2008;2(4):e228.

28. Khalil H, Santana R, de Oliveira D, Palma F, Lustosa R, Eyre MT, et al. Poverty, sanitation, and Leptospira transmission pathways in residents from four Brazilian slums. PLoS Negl Trop Dis. 2021;15(3):e0009256.

29. Costa F, Ribeiro GS, Felzemburgh RD, Santos N, Reis RB, Santos AC, et al. Influence of household rat infestation on leptospira transmission in the urban slum environment. PLoS Negl Trop Dis. 2014;8(12):e3338.

30. Felzemburgh RD, Ribeiro GS, Costa F, Reis RB, Hagan JE, Melendez AX, et al. Prospective study of leptospirosis transmission in an urban slum community: role of poor environment in repeated exposures to the Leptospira agent. PLoS Negl Trop Dis. 2014;8(5):e2927.

31. Linhares GP, Zequinao T, Buso GM, Cruz JAW, Tuon FF. Burden of leptospirosis in Brazil in the last decade. Rev Saude Publica. 2024;58:53.

32. Teles AJ, Bohm BC, Silva SCM, Bruhn FRP. Socio-geographical factors and vulnerability to leptospirosis in South Brazil. BMC Public Health. 2023;23(1):1311.

33. Hagan JE, Moraga P, Costa F, Capian N, Ribeiro GS, Wunder EA, et al. Spatiotemporal Determinants of Urban Leptospirosis Transmission: Four-Year Prospective Cohort Study of Slum Residents in Brazil. PLoS Negl Trop Dis. 2016;10(1):e0004275.

34. Azevedo TS, Nisa S, Littlejohn S, Muylaert RL. Leptospirosis in Campinas, Brazil: The interplay between drainage, impermeable areas, and social vulnerability. PLoS Negl Trop Dis. 2025;19(9):e0013560.

35. KP H, GA S, JS C, D dO, N N, JC L, et al. Influence of Rainfall on Leptospira Infection and Disease in a Tropical Urban Setting, Brazil - PubMed. Emerging infectious diseases. 2020 Feb;26(2).

36. Teles RdCCC, de Souza Silva A, Magalhães LS, Barros AP, de Araújo KCGM, de Santana Campos RN. ANÁLISE TEMPORAL E ESPACIAL DA LEPTOSPIROSE NO ESTADO DE SERGIPE SOB PERSPECTIVA DA SAÚDE ÚNICA. Revista Sergipana de Saúde Pública. 2025;4(1).

37. Martins MHDM, Spink MJP. Human leptospirosis as a doubly neglected disease in Brazil. Cien Saude Colet. 2020;25(3):919–28.

38. Branco LCdAC, Sanfins BZ, Souza FE, Campos AC, de Moraes VLS, de Andrade Ruela G. Perfil epidemiológico da leptospirose no Brasil de 2018 a 2023. Revista de Epidemiologia e Controle de Infecção. 2025;15(3).

39. Bezerra AP, de Souza A, de Medeiros ES, da Silva EB, de Oliveira Júnior JF. Modeling leptospirosis incidence in the northeast of Brazil using probability distributions and climatic factors: Insights for public health. Geosystems and Geoenvironment. 2025:100458.

40. Lau CL, Smythe LD, Craig SB, Weinstein P. Climate change, flooding, urbanisation and leptospirosis: fuelling the fire? Trans R Soc Trop Med Hyg. 2010;104(10):631–8.

41. Grimm AM, Ferraz SE, Gomes J. Precipitation anomalies in southern Brazil associated with El Niño and La Niña events. Journal of climate. 1998;11(11):2863–80.

42. Marengo J. Riscos das Mudanças Climáticas no Brasil: Análise Conjunta Brasil-Reino Unido sobre os Impac tos das Mudanças Climática s e do Desmatamento na Amazônia. 2011.

43. Coelho MS, Massad E. The impact of climate on Leptospirosis in São Paulo, Brazil. Int J Biometeorol. 2012;56(2):233–41.

44. Ghizzo Filho J, Nazário NO, Freitas PF, Pinto GdA, Schlindwein AD. Temporal analysis of the relationship between leptospirosis, rainfall levels and seasonality, Santa Catarina, Brazil, 2005-2015. Revista do Instituto de Medicina Tropical de São Paulo. 2018;60:e39.

45. Silva AEP, Neto FC, de Souza Conceição GM. Leptospirosis and its spatial and temporal relations with natural disasters in six municipalities of Santa Catarina, Brazil, from 2000 to 2016. Geospatial Health. 2020;15(2).

46. Hernandez CJ, Gunzel G, Ritter C, Bugs RCF, Rocha T, Fuller T, et al. Leptospirosis? An epidemiologic investigation following the historic 2024 floods in Rio Grande do Sul, Brazil. One Health. 2025:101146.

47. Ranieri T, Viegas da Silva E, Vallandro M, Oliveira Md, Barcellos R, Lenhardt R, et al. Leptospirosis Cases During the 2024 Catastrophic Flood in Rio Grande Do Sul, Brazil. Pathogens, 14 (4), 393. 2025.

48. de Albuquerque NF, Martins G, Medeiros L, Lilenbaum W, Ribeiro VMF. The role of capybaras as carriers of leptospires in periurban and rural areas in the western Amazon. Acta Trop. 2017;169:57–61.

49. Buffon FT, Pinheiro JAC, Castro HP, Barbosa FDADR, Mendonça RR, Gomes WR, et al. Enchente de 2015 no rio Acre: aquisição de dados e monitoramento. 2015.

50. da Silva SS, Brown F, de Oliveira Sampaio A, Silva ALC, dos Santos NCRS, Lima AC, et al. Amazon climate extremes: Increasing droughts and floods in Brazil’s state of Acre. Perspectives in Ecology and Conservation. 2023;21(4):311–7.

51. Melchior LAK, da Silva KRC, Silva AEP, Chiaravalloti-Neto F. Spatial, temporal, and space-time analysis of leptospirosis cases in Acre, 2001-2022. Rev Bras Epidemiol. 2024;27:e240063.

52. Governo do estado do Acre SdEdP. Plano Emergencial de Enfrentamento às Enchentes. 2024.

53. Epelboin L, Bourhy P, Le Turnier P, Schaub R, Mosnier E, Berlioz-Arthaud A, et al. La leptospirose en Guyane française et sur le bouclier des Guyanes. État des connaissances en 2016. Bulletin de la Société de pathologie exotique. 2017;110(3):165–79.

54. Correia LO, Padilha BM, Vasconcelos SM. [Methods for assessing the completeness of data in health information systems in Brazil: a systematic review]. Cien Saude Colet. 2014;19(11):4467–78.

